# Incubation period, serial interval, generation time and reproduction number of mpox clade I

**DOI:** 10.1101/2024.05.10.24307157

**Authors:** Valentina Marziano, Giorgio Guzzetta, Ira Longini, Stefano Merler

**Author notes:** joint senior authors (Ira Longini, Stefano Merler).

## Abstract

We estimate that the generation time of mpox clade I is distinctly longer than clade IIb and may depend on the transmission route (mean 17.5 days in households vs. 11.4 in hospitals). We estimate a mean reproduction number of 1.22-1.33 in the Democratic Republic of the Congo between August 2023-March 2024.

---

A large outbreak of mpox clade I (previously known as the Congo-Basin clade) has been ongoing in the Democratic Republic of the Congo (DRC) since the autumn of 2023 ^1^ and has caused, as of April 4, 2024, almost 20,000 cases and over 1,000 deaths (mostly among children under 15 years of age) across the whole country ^2^. With a case-fatality-ratio of up to 10% ^1^, infections with mpox clade I are significantly more deadly then clade IIb (formerly West African clade), which spread internationally since 2022, mainly through sexual contacts among men-who-have-sex-with-men (MSM) ^3^. The epidemiology of mpox clade I seems to be evolving: until recently, it was mainly of zoonotic origin, with only sporadic human-to-human transmission within households. Sexual transmission of mpox clade I has been observed during an outbreak investigation in Kwango Province ^4^ in March 2023. A new subclade has been recently identified in Kamituga, in the South Kivu Province ^5^, where extensive sexual transmission has been documented ^6,7^. Sustained human-to-human transmission, new routes of infections and genetic evolution raise important concerns about the potential risks of international spread, making it urgent to estimate critical epidemiological parameters specific to mpox clade I.

## This study

We leverage previously published outbreak investigation data to provide novel estimates of probability distribution functions for epidemiological parameters that are critical for monitoring and modelling the spread of mpox clade I.

We fitted three families of distributions (i.e., Weibull, gamma and log-normal (with a possible offset parameter)) to data from 15 cases with known incubation period (i.e., the time elapsed between the infection episode and symptom onset), reported in a recent review ^8^. The best estimate was to a Weibull distribution with an offset term of 4 days and mean 9.9 days (95% confidence interval, CI: 8.5-11.5 days) (Figure 1A).

**Figure 1.**
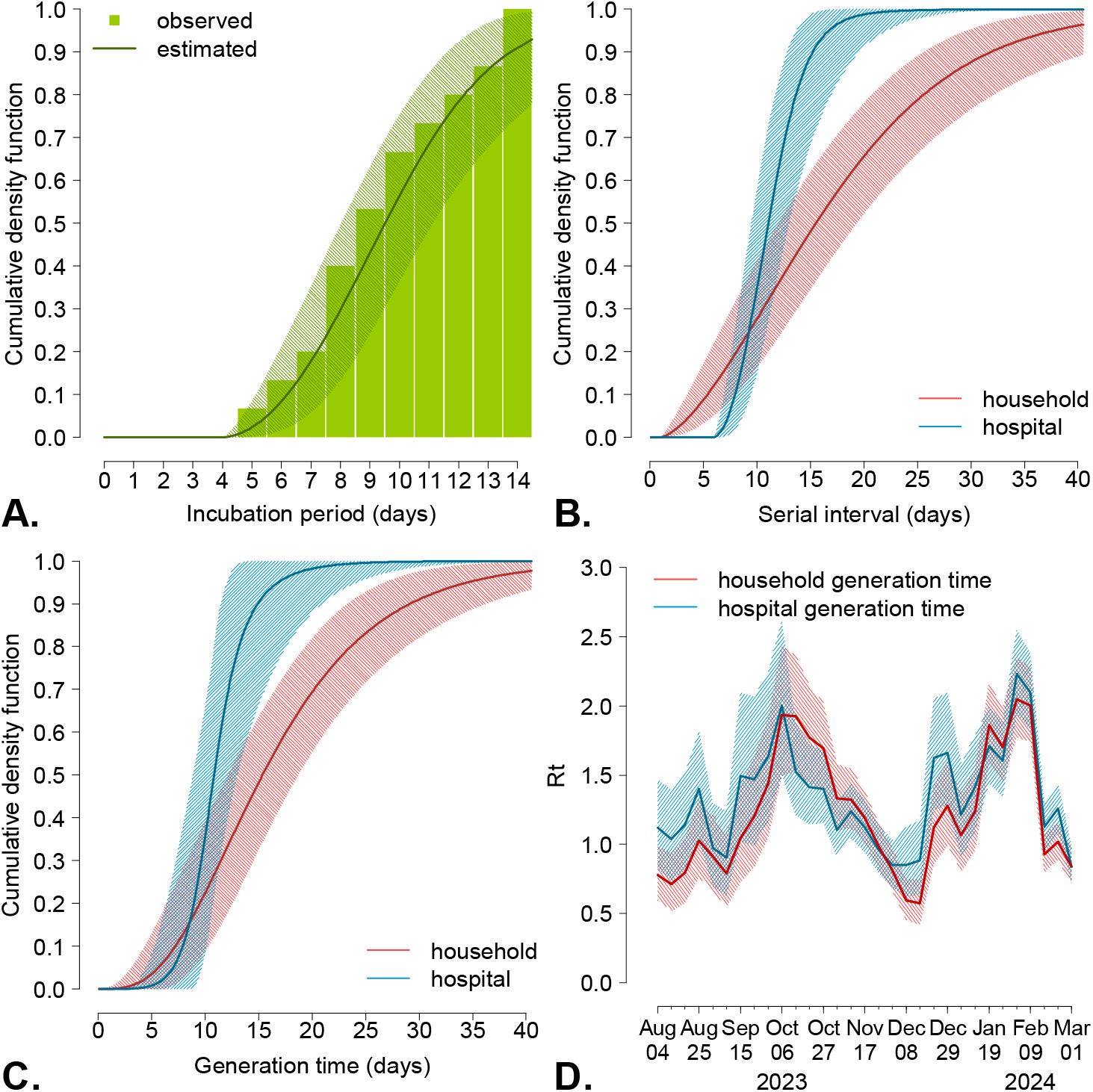
Estimates of key epidemiological parameters for mpox clade I. **A)** Cumulative density function of the incubation period, estimated from data on 15 cases reported in a published review ^8^. **B)** Cumulative density function of the serial interval, estimated from data on 32 transmission links identified within household outbreaks ^9,10^, and on data on 11 transmission links identified within a hospital outbreak ^11^. **C)** Cumulative density function of the estimated generation time, based on the same data reported for the serial interval and on estimates of the incubation period. **D)** Estimates of the time-varying reproduction number (Rt) in the Democratic Republic of Congo obtained from the time-series of reported cases^14^ and using the two estimates of the generation times.

We estimated two different distributions for the serial interval (i.e., the time elapsed between the symptom onset in an index case and in their secondary cases). The first was obtained by pooling together symptom onset dates for 32 infector-infectee transmission links from two household outbreaks in South Sudan, 2005 ^9^ (n=13) and in Central African Republic, 2021-2022 ^10^ (n=19). The second estimate was obtained from 11 infector-infectee transmission links from a hospital-associated outbreak in the Republic of the Congo, 2003 ^11^. In all three outbreaks, the chains of transmission were reconstructed through detailed epidemiological investigations identifying the most likely infector. We fitted the same three families of distributions, obtaining as an optimal estimate a Weibull with an offset term of 1 day and mean 17.5 days (95%CI: 14.1–21.5 days) for the household outbreaks and a Weibull with offset 6 days and mean 11.4 days (95%CI: 9.9-13.5 days) for the hospital outbreak data (Figure 1B).

The generation time (i.e. the time elapsed between the date of exposure of a confirmed case and those of their secondary cases) was estimated by considering the same data on infector-infectee pairs used for the estimate of the serial interval, following a Bayesian approach previously applied to mpox clade IIb ^12,13^. The generation time was assumed to be distributed as a gamma function with offset; parameters of the gamma and dates of exposure for all cases were estimated within a Markov chain Monte Carlo (MCMC) procedure. If the sampled date of exposure for the infectee was earlier than the date of symptom onset for the infector, this was considered an episode of pre-symptomatic transmission. The mean estimated generation time was 17.2 days (95%CI: 14.2-20.8 days) for household outbreaks and 11.3 days (95%CI: 9.5-13.6 days) for hospital outbreaks (Figure 1C), respectively. Pre-symptomatic transmission was likely negligible (i.e., about 7% of cases for household outbreaks and <1% of cases for hospital outbreaks) and parameter estimates obtained by not allowing pre-symptomatic transmission were virtually identical.

Estimates of the generation time were used to compute the net reproduction number (R_t_) in the DRC between early August 2023 and March 2024, based on the available time-series of country-wide weekly confirmed mpox cases ^14^ and on the renewal equation ^15–17^. Note that DRC has not reported mpox clade IIb cases ^2^ and, therefore, it is safe to assume that all mpox cases reported are due to clade I. R_t_ is defined as the average number of secondary cases per infectious individual at time t and represents a critical parameter to assess the transmissibility of an infection over time; when R_t_ <1, transmission is expected to fade out, whereas if R_t_ >1, the epidemic has the potential for further spread. We estimate a quite variable net reproduction number between August 4, 2023 and March 1, 2024 (Figure 1D), with mean 1.22 (range: 0.56-2.06) when considering the generation time estimated from household outbreaks and 1.33 (range: 0.84-2.21) when considering hospital outbreaks. Using provisional point estimates of the growth rate evaluated from phylodynamic analysis of the Kamituga outbreak ^5^ and our estimates of the generation time under the equation provided by Wallinga and Lipsitch ^15^ results in a local reproduction number for Kamituga of 1.39-1.61, higher than the national average. Full analysis details are given in the Technical Appendix.

## Discussion

The estimated mean incubation period of mpox clade I (95%CI: 8.5 - 11.5 days) was longer than most estimates reported for mpox clade IIb in a review by the WHO ^18^. Available data, although limited in sample size, point to a remarkable difference in serial intervals/generation times depending on the route of transmission, with household-associated cases having mean values ranging between 14 and 21 days (mean 17 days) and hospital-associated cases between 9 and 14 days (mean 11 days). Corresponding mean estimates for mpox clade IIb range between 5.6 and 9.4 days for serial intervals^18^ and about 12.5 days for generation times ^13^, suggesting a slower transmission dynamics of mpox clade I compared to IIb. We suggest a likely negligible occurrence of pre-symptomatic transmission, based on the estimated distribution of the infectious period and available data on serial intervals. This is again in contrast with mpox clade IIb, for which pre-symptomatic viral shedding and transmission has been demonstrated ^19,20^. As of today, there are no published data that allow the estimation of equivalent distributions for sexual transmission; furthermore, the distributions observed in the general community will likely depend on the relative contribution of each route of transmission.

The estimated reproduction number of the mpox clade I outbreak in DRC has been above the epidemic threshold at an average value of 1.2-1.3 since August 2023. The reproduction number estimated for a recently identified subclade which seems to have emerged in the South Kivu province of DRC ^5^ was even higher, at values of 1.4-1.6, possibly suggesting viral adaptation to human transmission; another possible interpretation is higher local contact rates due to sexual transmission ^7^ given the concentration of sex workers associated with the mining industry in the region ^2^.

We acknowledge that parameter estimations may be affected by small sample sizes and that the estimation of R_t_ may be distorted by time-varying diagnostic delays affecting the time series of cases by date of diagnosis.

These results show a distinct timing of mpox clade I epidemiology compared to mpox clade IIb, suggest the need for investigating the contribution of sexual transmission and associated serial intervals and generation times, and provide useful parameters for monitoring and modelling the transmission dynamics of mpox clade I. Finally, if not controlled with contact tracing and vaccination, this continued, relatively low level but persistent transmission of clade I mpox in the DRC could lead to further evolution of the virus towards higher person-to-person transmissibility and spread beyond the current geographic focus of transmission ^21^.

**Table 1.** Estimated parameters for the distributions of the incubation period, serial interval and generation time.

|  | Incubation<br>period | Serial interval<br>(households) | Serial interval<br>(hospital) | Generation time<br>(households) | Generation<br>time<br>(hospital) |
| --- | --- | --- | --- | --- | --- |
| Data source | Nolen et al.<sup>8</sup><br>N=15 | Formenty et al.<sup>9</sup><br>Besombes et al.<sup>10</sup><br>N=32 | Learned et<br>al.<sup>11</sup><br>N=11 | Formenty et al.<sup>9</sup><br>Besombes et al.<sup>10</sup><br>N=32 | Learned et<br>al.<sup>11</sup><br>N=11 |
| Distribution | Weibull | Weibull | Weibull | Gamma | Gamma |
| Offset median<br>(95%CI) (days) | 4<br>(fixed) | 1<br>(fixed) | 6<br>(fixed) | 1<br>(0-6) | 3<br>(0-8) |
| Shape mean<br>(95%CI) | 2.22<br>(1.38–3.22) | 1.63<br>(1.20–2.12) | 2.17<br>(1.29-3.19) | 3.19<br>(1.47-6.18) | 16.0<br>(1.46 – 76.34) |
| Scale mean<br>(95% CI) | 6.56<br>(5.01–8.44) | 18.29<br>(14.35–22.76) | 6.10<br>(4.38-8.38) | 5.38<br>(2.57-9.64) | 1.16<br>(0.13-4.00) |
| Mean<br>(95% CI) (days) | 9.9<br>(8.5–11.5) | 17.5<br>(14.1–21.5) | 11.4<br>(9.9-13.5) | 17.1<br>(14.0-20.6) | 11.3<br>(9.4-13.6) |

## Supporting information

Technical Appendix

## Data Availability

All data used in the present work are publicly available. References for each dataset used are reported in the main text.

## Acknowledgements

This research was supported by EU funding within the NextGenerationEU-MUR PNRR Extended Partnership Initiative on Emerging Infectious Diseases (project no. PE00000007, INF-ACT) received by SM. IL received funding from the Epistorm: Center for Advanced Epidemic Analytics and Predictive Modeling Technology (CDC, NU38FT000013). IL was partially supported by cooperative agreement CDC-RFA-FT-23-0069 from the CDC’s Center for Forecasting and Outbreak Analytics. The contents of this article are solely the responsibility of the authors and do not necessarily represent the official views of the Centers for Disease Control and Prevention.

