## Supplementary material for "Incubation period, serial interval, generation time and reproduction number of mpox clade I": Technical Appendix

*joint senior authors

**S1. Incubation period**

To estimate the distribution of the incubation period, we considered observations from 15 confirmed cases with known dates of exposure and symptom onset identified in (1) and reported in Table S1. We fitted three families of distributions (a gamma, a Weibull and a log-normal) assuming an offset of 4 days and using maximum likelihood estimation (MLE). The assumed value for the offset was based on the minimum observed incubation period of 5 days (Table S1). A Weibull distribution was selected based on the minimum value of the Bayesian information Criterion (BIC) score (Table S2 and Figure S1). The posterior distributions of the shape (k) and scale parameters ($\lambda$) were then estimated with a Monte Carlo Markov Chain (MCMC) procedure and Metropolis-Hastings sampling (2), as reported in the main text.

**Table S1.** Incubation periods (days) for 15 mpox cases with known dates of exposure and symptom onset (1).

| **ID case patient** | 1 | 2 | 3 | 4 | 5 | 6 | 7 | 8 | 9 | 10 | 11 | 12 | 13 | 14 | 15 |
|---|---|---|---|---|---|---|---|---|---|---|---|---|---|---|---|
| **Incubation periods (days)** | 5 | 6 | 7 | 8 | 8 | 8 | 9 | 9 | 10 | 10 | 11 | 12 | 13 | 14 | 14 |

**Table S2.** BIC score as obtained from MLE fit of the three families of distributions against data reported in Table S1. The best-fitting value of the three scores is highlighted in bold.

| **Distribution** | **BIC** |
|---|---|
| Gamma | 77.9 |
| Weibull | **76.8** |
| Log-normal | 80.2 |

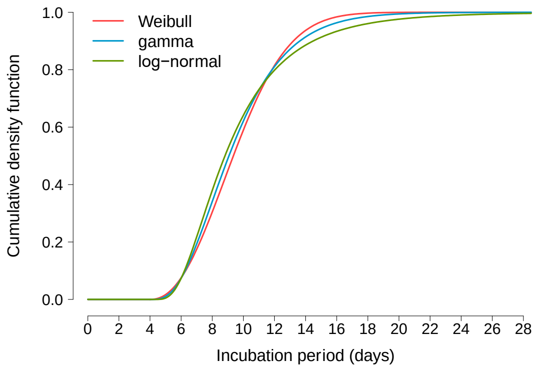


**Figure S1. Incubation period of mpox clade I.** Comparison between the cumulative probability distribution functions of the incubation period obtained from MLE for the three families of distributions.

**S2. Serial interval**

The serial interval is defined as the difference between the date of symptom onset of an infector and those of their infectees. We estimated the serial interval distribution using two different sets of infector-infectee pairs for which the dates of symptom onset were known. The first dataset consists of 32 pairs obtained by pooling together data from two household outbreaks in South Sudan, 2005 (3) (n=13) and in Central African Republic, 2021-2022 (4) (n=19) (Table S3); the second dataset consists of 11 pairs from a hospital-associated outbreak in the Republic of the Congo, 2003 (5) (Table S4).

**Table S3.** Information on infector-infectee pairs from household outbreaks.

| ID pair | ID infector | ID infected | Symptom onset infector | Symptom onset infected | Observed serial interval | Source |
|---|---|---|---|---|---|---|
| 1 | B1 | B2 | 06/11/2021 | 24/11/2021 | 18 | Besombes et al. (4) |
| 2 | B1 | B3 | 06/11/2021 | 27/11/2021 | 21 | Besombes et al. (4) |
| 3 | B1 | B4 | 06/11/2021 | 30/11/2021 | 24 | Besombes et al. (4) |
| 4 | B1 | B5 | 06/11/2021 | 08/11/2021 | 2 | Besombes et al. (4) |
| 5 | B6 | B7 | 07/11/2021 | 02/12/2021 | 25 | Besombes et al. (4) |
| 6 | B6 | B8 | 07/11/2021 | 06/12/2021 | 29 | Besombes et al. (4) |
| 7 | B6 | B9 | 07/11/2021 | 13/12/2021 | 36 | Besombes et al. (4) |
| 8 | B4 | B10 | 30/11/2021 | 02/12/2021 | 2 | Besombes et al. (4) |
| 9 | B10 | B11 | 02/12/2021 | 04/01/2022 | 33 | Besombes et al. (4) |
| 10 | B7 | B12 | 02/12/2021 | 08/12/2021 | 6 | Besombes et al. (4) |
| 11 | B9 | B13 | 13/12/2021 | 16/12/2021 | 3 | Besombes et al. (4) |
| 12 | B9 | B14 | 13/12/2021 | 19/12/2021 | 6 | Besombes et al. (4) |
| 13 | B12 | B15 | 08/12/2021 | 20/12/2021 | 12 | Besombes et al. (4) |
| 14 | B12 | B16 | 08/12/2021 | 23/12/2021 | 15 | Besombes et al. (4) |
| 15 | B12 | B17 | 08/12/2021 | 28/12/2021 | 20 | Besombes et al. (4) |
| 16 | B12 | B18 | 08/12/2021 | 01/01/2022 | 24 | Besombes et al. (4) |
| 17 | B12 | B19 | 08/12/2021 | 04/01/2022 | 27 | Besombes et al. (4) |
| 18 | B12 | B20 | 08/12/2021 | 12/01/2022 | 35 | Besombes et al. (4) |
| 19 | B12 | B21 | 08/12/2021 | 12/01/2022 | 35 | Besombes et al. (4) |
| 20 | F1 | F2 | 19/09/2005 | 30/09/2005 | 11 | Formenty et al. (3) |
| 21 | F2 | F3 | 30/09/2005 | 08/10/2005 | 8 | Formenty et al. (3) |
| 22 | F2 | F4 | 30/09/2005 | 08/10/2005 | 8 | Formenty et al. (3) |
| 23 | F2 | F5 | 30/09/2005 | 16/10/2005 | 16 | Formenty et al. (3) |
| 24 | F2 | F6 | 30/09/2005 | 16/10/2005 | 16 | Formenty et al. (3) |
| 25 | F5 | F7 | 16/10/2005 | 30/10/2005 | 14 | Formenty et al. (3) |
| 26 | F5 | F8 | 16/10/2005 | 03/11/2005 | 18 | Formenty et al. (3) |
| 27 | F6 | F9 | 16/10/2005 | 30/10/2005 | 14 | Formenty et al. (3) |
| 28 | F10 | F11 | 27/10/2005 | 05/11/2005 | 9 | Formenty et al. (3) |
| 29 | F12 | F13 | 01/12/2005 | 15/12/2005 | 14 | Formenty et al. (3) |
| 30 | F12 | F14 | 01/12/2005 | 15/12/2005 | 14 | Formenty et al. (3) |
| 31 | F15 | F16 | 24/10/2005 | 08/11/2005 | 15 | Formenty et al. (3) |
| 32 | F15 | F17 | 24/10/2005 | 08/11/2005 | 15 | Formenty et al. (3) |

**Table S4.** Information on infector-infectee pairs from hospital associated outbreak.

| ID pair | ID infector | ID infected | Symptom onset infector | Symptom onset infected | Serial interval | Source |
|---|---|---|---|---|---|---|
| 1 | L1 | L2 | 15/04/2003 | 28/04/2003 | 13 | Learned et al. (5) |
| 2 | L2 | L3 | 28/04/2003 | 08/05/2003 | 10 | Learned et al. (5) |
| 3 | L3 | L4 | 08/05/2003 | 18/05/2003 | 10 | Learned et al. (5) |
| 4 | L3 | L5 | 08/05/2003 | 18/05/2003 | 10 | Learned et al. (5) |
| 5 | L4/L5 | L6 | 18/05/2003 | 28/05/2003 | 10 | Learned et al. (5) |
| 6 | L4/L5 | L7 | 18/05/2003 | 30/05/2003 | 12 | Learned et al. (5) |
| 7 | L6 | L8 | 28/05/2003 | 05/06/2003 | 8 | Learned et al. (5) |
| 8 | L6 | L9 | 28/05/2003 | 05/06/2003 | 8 | Learned et al. (5) |
| 9 | L7 | L10 | 30/05/2003 | 10/06/2003 | 11 | Learned et al. (5) |
| 10 | L10 | L11 | 10/06/2003 | 23/06/2003 | 13 | Learned et al. (5) |
| 11 | L8 | L12 | 05/06/2003 | 22/06/2003 | 17 | Learned et al. (5) |

We used MLE to fit the same three families of distributions (gamma, Weibull and log-normal) to the observed serial intervals, assuming an offset of 1 day for the household outbreaks and of 6 days for the hospital outbreak, based on the minimum observed serial intervals (BIC scores reported in Table S5). For household outbreaks, the best-fitting distribution was the Weibull, while for hospital outbreak the log-normal was marginally better than the gamma and Weibull. Given the small differences among the distributions reported in Table S5 and shown in Figure S2, to maintain uniformity we selected the Weibull distribution for the hospital dataset as well. For both the household and hospital dataset, we estimated the posterior distributions of the shape (k) and scale parameters ($\lambda$) of the Weibull distribution, based on MCMC and Metropolis-Hastings sampling (2). The posterior distributions of the serial interval obtained for households and hospital outbreaks, respectively, are shown in Figure S3.

**Table S5.** BIC score obtained from the MLE fit of the three families of distributions to serial interval data from the household and hospital outbreaks. The best-fitting values are highlighted in bold.

| | **BIC** | |
|---|---|---|
| **Distribution** | **Household (n=32)** | **Hospital (n=11)** |
| **Gamma** | 241.5 | 53.64 |
| **Weibull** | **239.2** | 54.38 |
| **Log-normal** | 250.5 | **53.61** |

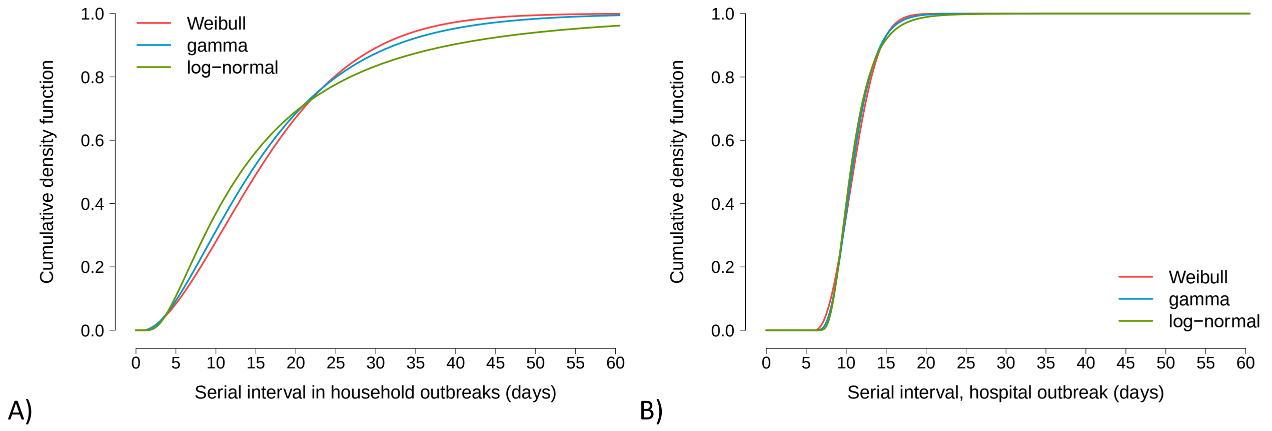


**Figure S2.** Comparison between the probability distribution functions of the serial interval obtained from MLE for the three families of distributions. A) Household outbreaks; B) hospital outbreak.

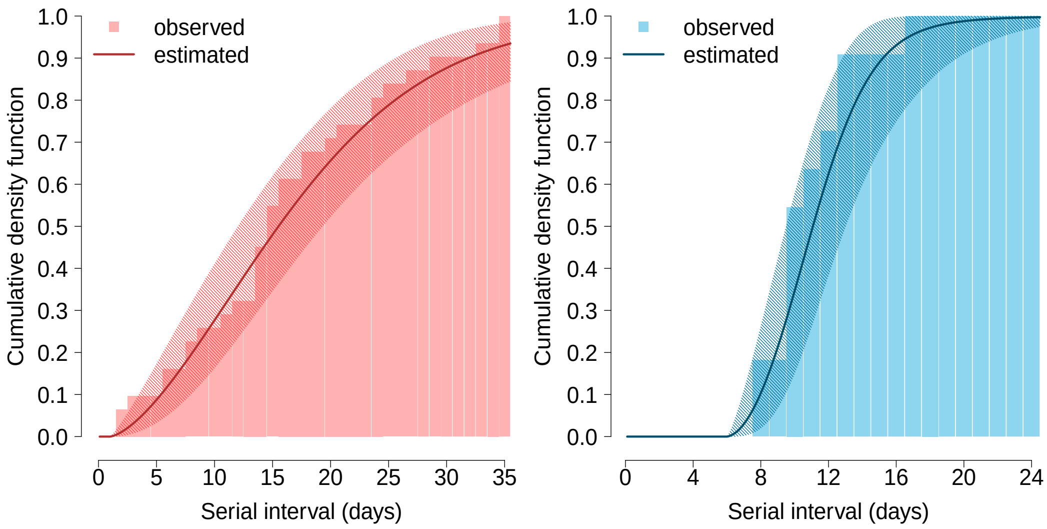


**Figure S3. Serial interval of mpox clade I.** Left: cumulative density function of the serial interval, estimated from data on 32 infector-infectee pairs reported in the literature in household outbreaks (3,4). Right: cumulative density function of the serial interval, estimated from data on 11 infector-infectee pairs reported in the literature in a hospital outbreak (5).

### S3. Generation time

The generation time is defined as the difference between the date of infection of an infector and those of their infectees. We estimated two generation time distributions, one for household outbreaks and one for the hospital outbreak, based on the same infector-infectee transmission pairs considered for the serial interval (Table S3 and S4 respectively). For each dataset, we estimated the posterior distributions of the offset ($\theta$), the shape (k) and scale ($\lambda$) parameters of a gamma distribution, based on an MCMC procedure with Metropolis-Hastings sampling, where the infectious dates are considered as dummy parameters and the incubation period associated with them is included in the definition of the likelihood function (2,6).

*S3.1 Sensitivity analysis without pre-symptomatic transmission*

In the main analysis, infection dates are sampled freely, therefore, the sampled date of exposure of infectees may precede the date of symptom onset of their infector, representing a potential pre-symptomatic transmission. Since it is not known whether asymptomatic transmission is possible for mpox clade I, we run a sensitivity analysis where we constrained the date of infection for the secondary cases to be always greater or equal to the date of symptom onset of the infector, i.e., if this condition is violated, the sample will be rejected. The estimated parameters for the generation time distributions are reported in Table S6 and are similar to the baseline analysis. The cumulative density functions of the estimated generation times are shown in Figure S4.

**Table S4.** Estimated parameters for the distributions of the generation time assuming that pre-symptomatic transmission is not allowed.

| | **Generation time (households)** | **Generation time (hospital)** |
|---|---|---|
| **Data source** | Formenty et al. (3)<br>Besombes et al. (4) | Learned et al. (5) |
| **Distribution** | Gamma | Gamma |
| **Offset median (95%CI) (days)** | 2 (0-7) | 3 (0-8) |
| **Shape mean (95%CI)** | 3.41 (1.30-7.01) | 18.33 (1.47 –96.51) |
| **Scale mean (95% CI)** | 4.95 (2.23-9.61) | 1.06 (0.1-3.64) |
| **Mean (95% CI) (days)** | 17.2 (14.2-20.8) | 11.3 (9.5-13.6) |

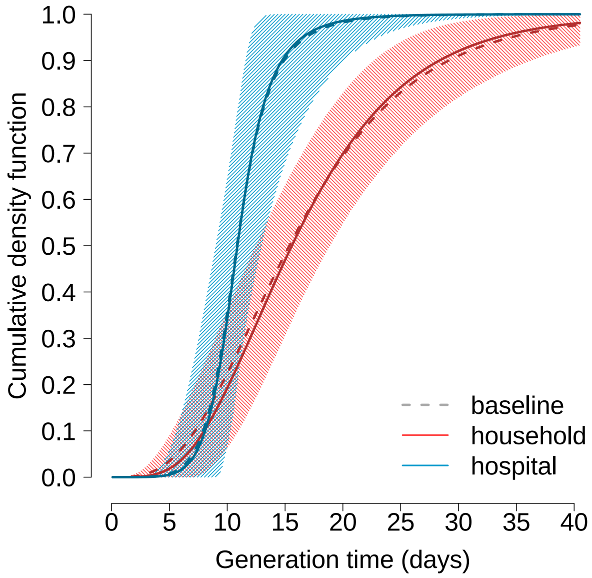


**Figure S4.** Cumulative density functions of the estimated generation times in household and hospital outbreaks as obtained by assuming that pre-symptomatic transmission is not allowed. The corresponding mean distributions obtained in the baseline analysis are reported with a dashed line for comparison.

**S4. Reproduction number**

Temporal variations in the transmissibility of a pathogen can be monitored through the net reproduction number, $R_t$, defined as the average number of secondary cases per infectious individual at time t. We estimate $R_t$ from weekly confirmed mpox cases in the Democratic Republic of the Congo between August 4, 2023 and March 1, 2024 (7) (Figure S5) by applying a commonly used statistical method based on the renewal equation (8,9) and on the above-estimated distribution of the generation time. More specifically, the posterior distribution of $R_t$ was estimated by applying MCMC to the following likelihood function:

$$\mathcal{L} = \prod_{t=1}^{T} P\left( C(t) - I(t);\ R_t \sum_{s=1}^{T} \varphi(s) C(t-s) \right)$$

Where:

- $P(k; \lambda)$ is the probability mass function of a Poisson distribution (i.e., the probability of observing k events if these events occur with rate $\lambda$).
- $C(t)$ is the total weekly number of new cases confirmed at week t;
- $I(t)$ is the total weekly number of new cases that are not locally transmitted (imported from another geographic setting or from the animal reservoir);
- $R_t$ is the net reproduction number at time t to be estimated;
- $\varphi(s)$ is the probability mass function of the generation time discretized by week, evaluated at week s.

In absence of other information, we considered only one imported case at the beginning of the time series.

Estimates of $R_t$ obtained when considering the generation time distributions estimated in the sensitivity analysis, i.e. where pre-symptomatic transmission is not considered, are shown in Figure S6 and compared to the ones obtained in the main analysis in Table S5.

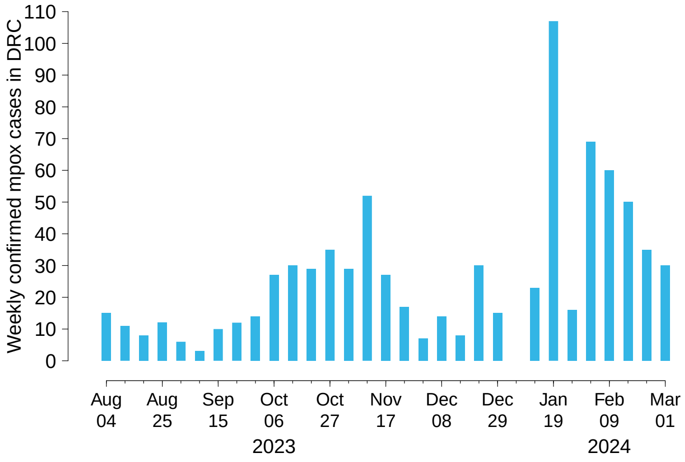


**Figure S5.** Time-series of weekly mpox confirmed cases in the Democratic Republic of the Congo (7).

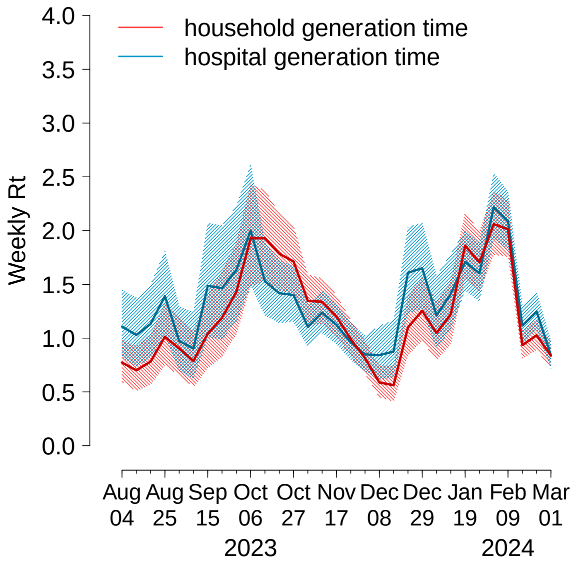


**Figure S6**. Estimates of the time-varying reproduction number ($R_t$) in the Democratic Republic of the Congo obtained from the time-series of reported cases and using the two estimates of the generation times (in household and hospital outbreaks), obtained in a sensitivity analysis where pre-symptomatic transmission is not allowed.

**Table S5.** Average value and range over the period August 4, 2023 – March 1, 2024 of the mean $R_t$ estimate.

| | **Generation time (households)** | **Generation time (hospital)** |
|---|---|---|
| **Pre-symptomatic transmission allowed (main analysis)** | 1.22 (0.56-2.06) | 1.33 (0.84-2.21) |
| **No pre-symptomatic transmission allowed (sensitivity analysis)** | 1.22 (0.57-2.05) | 1.33 (0.84-2.23) |

Finally, to compute the reproduction number R of an ongoing outbreak in Kamituga associated to a new subclade of mpox I (10), we applied the following formula that allows us to compute R from an estimate of the exponential growth rate *r* and of the generation time distribution $\varphi(t)$ (11):

$$R = \int_0^\infty \frac{1}{\varphi(t)e^{-rt}} dt$$

An exponential growth rate of $r = 10.8$ per year has been estimated from phylodynamic analyses (9); applying our estimates for the generation time distributions, we obtain results presented in Table S6.

**Table S6.** Point estimates of the reproduction number for the Kamituga outbreak.

| | Generation time (households) | Generation time (hospital) |
|---|---|---|
| **Pre-symptomatic transmission allowed (main analysis)** | 1.61 | 1.39 |
| **No pre-symptomatic transmission allowed (sensitivity analysis)** | 1.62 | 1.39 |
